# Evaluation of Ultrafast Wave-CAIPI 3D FLAIR in the Visualization and Volumetric Estimation of Cerebral White Matter Lesions

**DOI:** 10.1101/2021.01.10.21249348

**Authors:** Chanon Ngamsombat, Augusto Lio M. Gonçalves Filho, M. Gabriela Figueiro Longo, Stephen F. Cauley, Kawin Setsompop, John E. Kirsch, Qiyuan Tian, Qiuyun Fan, Daniel Polak, Wei Liu, Wei-Ching Lo, R. Gilberto González, Pamela W. Schaefer, Otto Rapalino, John Conklin, Susie Y. Huang

## Abstract

**BACKGROUND AND PURPOSE:** To evaluate an ultrafast 3D-FLAIR sequence using Wave-CAIPI encoding (Wave-FLAIR) compared to standard 3D-FLAIR in the visualization and volumetric estimation of cerebral white matter lesions in a clinical setting.

**MATERIALS AND METHODS:** 42 consecutive patients underwent 3T brain MRI including standard 3D-FLAIR (acceleration factor R=2, scan time TA=7:15 minutes) and resolution-matched ultrafast Wave-FLAIR sequences (R=6, TA=2:45 minutes for the 20-ch coil; R=9, TA=1:50 minutes for the 32-ch coil) as part of clinical evaluation for demyelinating disease. Automated segmentation of cerebral white matter lesions was performed using the Lesion Segmentation Tool in SPM. Student’s t-test, intra-class correlation coefficient (ICC), relative lesion volume difference (LVD) and Dice similarity coefficients (DSC) were used to compare volumetric measurements between sequences. Two blinded neuroradiologists evaluated the visualization of white matter lesions, artifact and overall diagnostic quality using a predefined 5-point scale.

**RESULTS:** Standard and Wave-FLAIR sequences showed excellent agreement of lesion volumes with an ICC of 0.99 and DSC of 0.97±0.05 (range 0.84 to 0.99). Wave-FLAIR was non-inferior to standard-FLAIR for visualization of lesions and motion. The diagnostic quality for Wave-FLAIR was slightly greater than standard-FLAIR for infratentorial lesions (p<0.001), and there was less pulsation artifact on Wave-FLAIR compared to standard FLAIR (p<0.001).

**CONCLUSIONS:** Ultrafast Wave-FLAIR provides superior visualization of infratentorial lesions while preserving overall diagnostic quality and yields comparable white matter lesion volumes to those estimated using standard-FLAIR. The availability of ultrafast Wave-FLAIR may facilitate the greater use of 3D-FLAIR sequences in the evaluation of patients with suspected demyelinating disease.

## Introduction

White matter lesions secondary to demyelination in multiple sclerosis (MS) and related disorders typically present with high T2 signal and are best evaluated with fluid attenuated inversion recovery (FLAIR) imaging, the standard sequence for cerebral white matter lesion detection. FLAIR is a T2-weighted sequence with nulling of the cerebrospinal fluid signal, which increases the contrast between lesions and CSF/cerebral sulci and ventricles and improves white matter lesion detection and analysis.^1^

Quantification of cerebral white matter lesion volume has become increasingly feasible for routine clinical evaluation and use in clinical trials of MS therapies due to the availability of automated segmentation tools and three-dimensional fast spin echo fluid-attenuated inversion recovery (3D FSE FLAIR) sequences, which delineate cerebral white matter lesions at high isotropic resolution. Lesion Segmentation Tool (LST), a promising tool for automated segmentation of T2 hyperintense lesions on FLAIR images, was developed for the quantification of MS lesion volumes and has been shown to have good agreement with manual segmentation by expert reviewers.^2-7^ However, the high-resolution 3D FLAIR images required as input for this tool suffer from long acquisition times, which has limited the widespread use of automated lesion segmentation in clinical practice.

Wave–controlled aliasing in parallel imaging (CAIPI) is a recently developed fast acquisition technology that synergistically combines and extends two controlled aliasing approaches, 2D-CAIPI and bunch phase encoding (BPE),^8^ to achieve controlled aliasing in all three spatial directions (x, y, z). By taking full advantage of the 3D coil sensitivity information, Wave-CAIPI offers high acceleration factors with negligible artifacts and g-factor penalty.^9, 10^ 3D FLAIR acquired with Wave-CAIPI cuts the scan time down by more than half, which may facilitate the broader clinical application of 3D FLAIR in the evaluation of white matter diseases such as MS.

The goal of this study was to evaluate an ultrafast Wave-CAIPI 3D FLAIR sequence (Wave-FLAIR)^11, 12^ acquired in less than half the time as standard 3D FLAIR for quantitative and qualitative analyses of cerebral white matter lesions.

## Methods

### Subjects and study design

This study was approved by the IRB and was HIPAA compliant. A prospective comparative study was performed at a single institution from April 2019 to March 2020. Forty-two consecutive patients undergoing brain MRI as part of routine clinical work-up and/or surveillance for multiple sclerosis (MS) and other white matter diseases were enrolled.

### Data acquisition

MRI scans were performed on one of two clinical 3T MR scanners (MAGNETOM Prisma, Siemens Healthcare, Erlangen, Germany) using 20- or 32-channel multi-array receiver coils, depending on the fit and comfort of the patient. Each scan included a standard 3D Sampling Perfection with Application optimized Contrasts by using different flip angle Evolutions (SPACE) FLAIR sequence (acceleration factor R=2, scan time TA=7:15 min) and resolution-matched ultrafast 3D Wave SPACE-FLAIR (R=6, TA=2.45 min for the 20-ch coil and R=9, TA=1:50 min for the 32-ch coil) sequences. The order of the Wave and standard-FLAIR sequences was reversed halfway through the study period in order to minimize any potential bias due to order of acquisition. Detailed acquisition parameters for the standard and Wave SPACE-FLAIR sequences are shown in Table 1.

**Table 1.** Acquisition parameters for Standard and Wave SPACE-FLAIR sequences

| Parameters |  |  | Standard | Wave SPACE-<br>FLAIR |
| --- | --- | --- | --- | --- |
| FOV read (mm) |  |  | 256 x 256 | 256 x 256 |
| FOV phase (%) |  |  | 100 | 100 |
| Matrix size |  |  | 256 x 256 | 256 x 256 |
| Slice thickness (mm) |  |  | 1 | 1 |
| TR/TE/TI (msec) |  |  | 5000/390/1800 | 5000/392/1800 |
| Acceleration factor | 20-ch |  | 2 | 6 |
|  | 32-ch |  | 2 | 9 |
| Bandwidth (Hz/pz) |  |  | 750 | 650 |
| Scan time (sec) | 20-ch |  | 7:15 min | 2:45 min |
|  | 32-ch |  | 7:15 min | 1:50 min |

### White matter lesion analysis

#### Quantitative analysis

Cerebral white matter lesions were segmented using the lesion prediction algorithm (LPA) implemented in the Lesion Segmentation Tool (LST) toolbox version 2.0.15 (www.statistical-modelling.de/lst.html) in the SPM^2^. Lesion probability maps generated by LPA from the standard and Wave-FLAIR sequences were compared using the longitudinal pipeline in LST. Binarized lesion maps were created based on the lesion probability maps derived from standard and Wave-FLAIR sequences using default threshold values set by LST for all subjects. Lesions in each brain regions including periventricular, juxtacortical, infratentorial, deep white matter, subcortical white matter and deep gray matter were identified and labeled by a neuroradiologist blinded to sequence type and order using the Island Tools Editor in 3DSlicer version 4.10.2 (https://www.slicer.org/) for further analyses. Lesion volume and number were compared in each brain region between the standard and Wave-FLAIR images.

#### Qualitative analysis

Two neuroradiologists (J.C. and A.L.G., 8 years of experience each) blinded to sequence type performed a head-to-head comparison of the images. A predefined 5-point scale was used for grading white matter lesions in the locations specified in the McDonald criteria (i.e., periventricular, juxtacortical and infratentorial locations)^13^ and other locations including subcortical white matter, deep white matter and deep gray matter. Other variables that were evaluated included motion, pulsation artifact, noise and overall diagnostic quality (Supplementary Table 1). All images were evaluated in a randomized and blinded fashion. A third reader adjudicated the discrepancies (S.Y.H., 10 years of experience).

#### Statistical analysis

All statistical calculations were all performed in MATLAB software version 9.4 (MathWorks, Natick, Massachusetts) and R statistical and computing software, Version 3.4.3 (http://www.r-project.org/). Student’s t-test was used to compare lesion volumes and lesion numbers between the standard and Wave sequences. The correlation between standard and Wave-FLAIR lesion volumes and lesion numbers were assessed using Pearson’s correlation coefficients. Two-way random intra-class correlation coefficient (ICC) for absolute agreement and for consistency^14^ were calculated to determine volumetric accuracy, with a higher ICC signifying higher inter-measurement agreement.^5, 7, 14, 15^ Relative lesion volume difference (LVD)^16^ was also used to compare standard and Wave-FLAIR lesion volumes, defined as LVD = (total lesion volume on Wave – total lesion volume on standard)/total lesion volume on standard. Dice similarity coefficients (DSC) were used to compare quantitative volumetric measurements between sequences.^3, 5, 7^ The Dice similarity coefficient of the standard and Wave images was expressed as dice(Standard,Wave) = 2∗ |intersection(Standard,Wave)| / (|Standard| + |Wave|). DSC measures have values between 0 and 1 with higher values indicating better agreement.^17^

In the head-to-head qualitative analysis of the standard versus Wave-FLAIR images, we tested for noninferiority of Wave compared to Standard-FLAIR^18^ using a noninferiority margin (Δ) of 15% as previously established.^19^ The null hypothesis (H_0_) was that the proportion of cases in which standard-FLAIR was preferred over Wave-FLAIR was >15%. We used the z statistic to calculate the probability of the standard-FLAIR being preferred over the Wave-FLAIR sequence in >15% of cases (H_0_ > Δ), with a type I error rate (α) of 0.05. The required sample size was estimated as described^20^ for a single proportion (the proportion of subjects in which visualization of lesions was preferred on standard over Wave-FLAIR), for a type I error rate (Δ) of 0.05, a power (1–Δ) of 0.90 and non-inferiority margin of 15%, a minimum of 24 cases was required. For all statistical analyses, corrections for multiple comparison were conducted based on the false discovery rate (FDR) adjustment with an FDR threshold of 0.05. The raw uncorrected p-values surviving FDR correction are reported here.

## Results

Forty-two adults participated in the comparative evaluation of the standard and Wave-FLAIR sequences. Demographic information on the study subjects, including age, sex and clinical indication for undergoing MRI, are shown in Supplementary Table 2. A total of 38 patients out of 42 (90.5%) had white matter lesions. 36 patients (85.7%) were scanned with the 20-channel coil. 22 cases were scanned with standard before Wave-FLAIR, while 20 were scanned with Wave before standard FLAIR.

**Table 2:** Comparison of lesion volume in different brain regions as assessed on standard and Wave-FLAIR images.

|  | **Standard** |  | **WAVE** |  | **t-test** | **ICC** | **LVD** | **DSC** |
| --- | --- | --- | --- | --- | --- | --- | --- | --- |
| **Brain regions** | Lesions<br>in all<br>patients<br>(mm<sup>3</sup>) | Mean (SD) | Lesions<br>in all<br>patients<br>(mm<sup>3</sup>) | Mean (SD) | p-value | ICC | Mean (SD) | Mean (SD) |
| **Whole brain** | 167800 | 4661.1<br>(13185) | 168130 | 4670.4<br>(13180) | 0.99 | 0.99 | 0.01 (0.05) | 0.97 (0.05) |
| **Periventricular** | 155789 | 4327.47<br>(13180.40) | 156121 | 4336.69<br>(13175.96) | 0.99 | 0.99 | 0.014 (0.06) | 0.99 (0.03) |
| **Juxtacortical** | 1892 | 52.56<br>(79.20) | 1881 | 52.25<br>(79.14) | 0.98 | 0.99 | -0.012 (0.05) | 0.91 (0.25) |
| **Infratentorial** | 617 | 17.14<br>(30.64) | 616 | 17.11<br>(30.50) | 0.99 | 0.99 | 0.014 (0.09) | 0.84 (0.32) |
| **Deep white matter** | 4654 | 129.28<br>(259) | 4650 | 129.17<br>(259.18) | 0.99 | 0.99 | 0.005 (0.06) | 0.98 (0.05) |
| **Subcortical white matter** | 1852 | 51.44<br>(104) | 1857 | 51.58<br>(103.42) | 0.99 | 0.99 | 0.014 (0.1) | 0.95 (0.19) |
| **Deep gray matter** | 2995 | 83.19<br>(416.66) | 3017 | 83.81<br>(416.73) | 0.99 | 0.99 | 0.02 (0.05) | 0.98(0.05) |

\* ICC = intra-class correlation coefficient; LVD = relative lesion volume difference; DSC = Dice similarity coefficient.

In this section, we first present the quantitative comparison of lesion volumes followed by the qualitative evaluation of image quality. Standard and Wave-FLAIR images were evaluated in each brain region using predefined evaluation metrics described in the Methods.

Six patients were excluded from the LST quantitative analysis as they had no detectable lesions and/or severe motion artifact resulting in failure of the automated LST processing stream. On the whole brain level, the standard and Wave-FLAIR sequences showed no significant difference in lesion volume (167800 vs 168130 mm^3^, p=0.99) or lesion number (520 vs 529, p=0.91) as segmented by LST. The ICC between standard and Wave-FLAIR was 0.99. The relative lesion volume difference (LVD) was 0.01± 0.05 [range = −0.012 – 0.02], and the Dice similarity coefficient (DSC) was 0.97 ± 0.05 [range = 0.84 – 0.99] (Table 2). For lesions in each brain region (Figure 1 and 2), there was excellent agreement between standard and Wave-FLAIR images for lesion volume and lesion number (Supplementary Figures 1 and 2), with no significant difference in lesion volumes (p>0.98) (Table 2) or lesion numbers (p>0.89) (Supplementary Table 3) as segmented by LST for each sequence. The LVD’s were very low for all brain regions (<2%), and Dice coefficient was greater than 0.9 between the two sequences for all brain regions including the periventricular, deep and subcortical white matter and deep gray matter, with the exception of the infratentorial white matter (DSC = 0.84) (Table 2).^∗^ ICC = intra-class correlation coefficient; LVD = relative lesion volume difference; DSC = Dice similarity coefficient.

**Figure 1.**
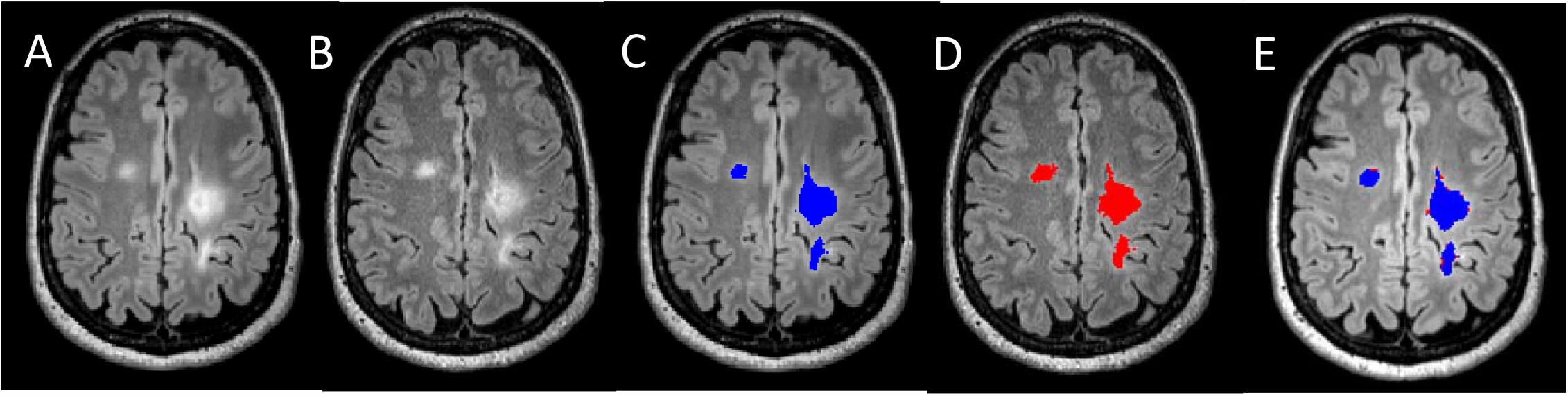
Comparison of standard and Wave SPACE-FLAIR images and lesion masks. (A) Standard SPACE-FLAIR image. (B) Wave SPACE-FLAIR image. (C) Lesion mask on standard SPACE-FLAIR image (blue). (D) Lesion mask on Wave SPACE-FLAIR image (red). (E) Lesion masks from standard and Wave SPACE-FLAIR images overlaid on the same image.

**Figure 2.**
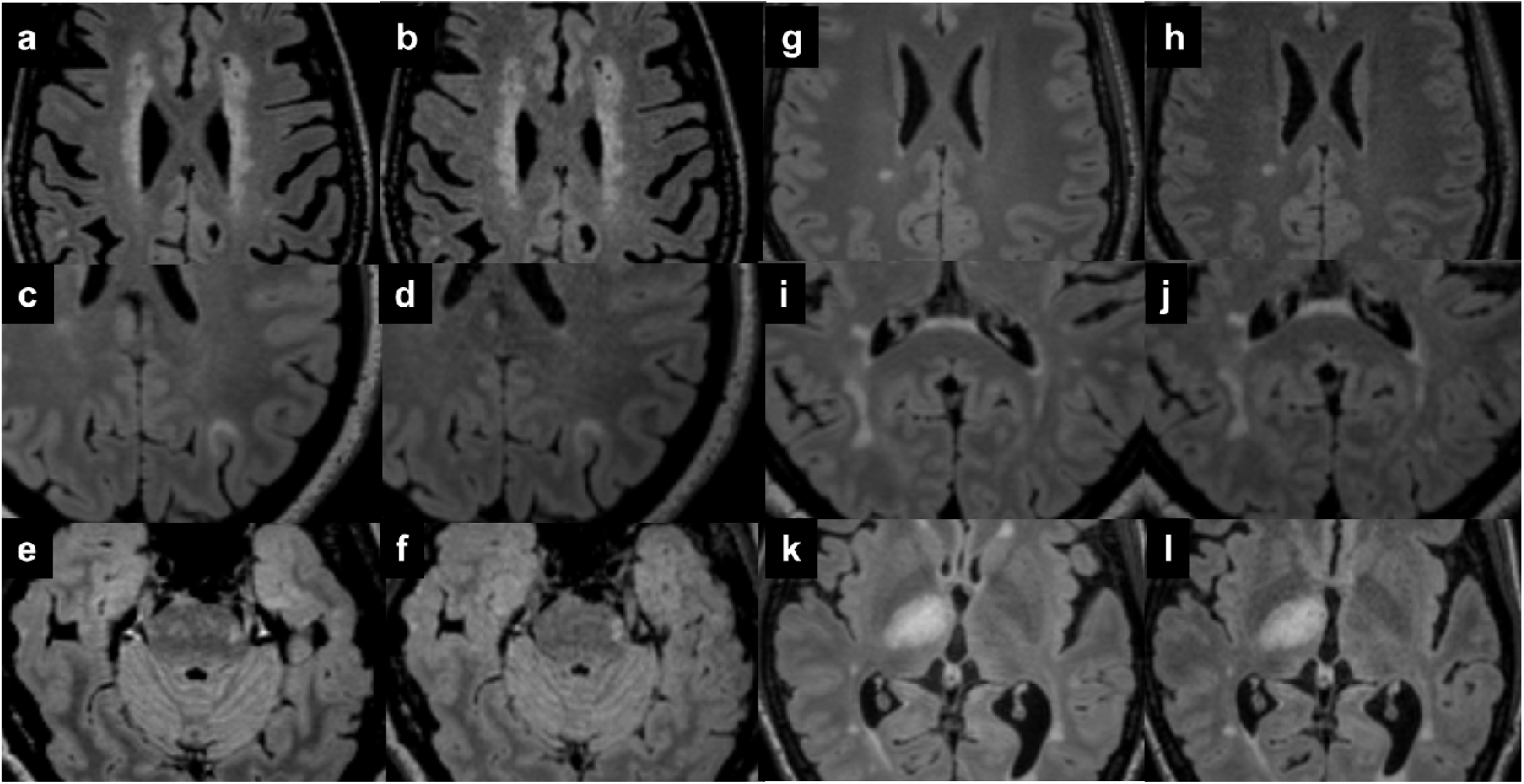
Comparison of MS lesions in different brain regions on standard and Wave-FLAIR images. The standard images (a, c, e, g, i, k) are on the left, and the Wave-FLAIR images (b, d, f, h, j, l) are on the right of each image pair. Lesion locations included the periventricular (a and b), juxtacortical (c and d), infratentorial (e and f), and deep white matter (g and h), subcortical white matter (i and j) and deep gray matter (k and l).

Wave-FLAIR was equivalent to standard FLAIR for the visualization of lesions in the subcortical and deep white matter and deep gray matter (p<0.001) and was noninferior to standard FLAIR in the visualization of periventricular (p<0.001), juxtacortical (p<0.006), and infratentorial lesions (p<0.001) (Figure 3). There was a slightly greater preference for Wave-FLAIR in the visualization of infratentorial lesions compared to the standard FLAIR sequence.

**Figure 3.**
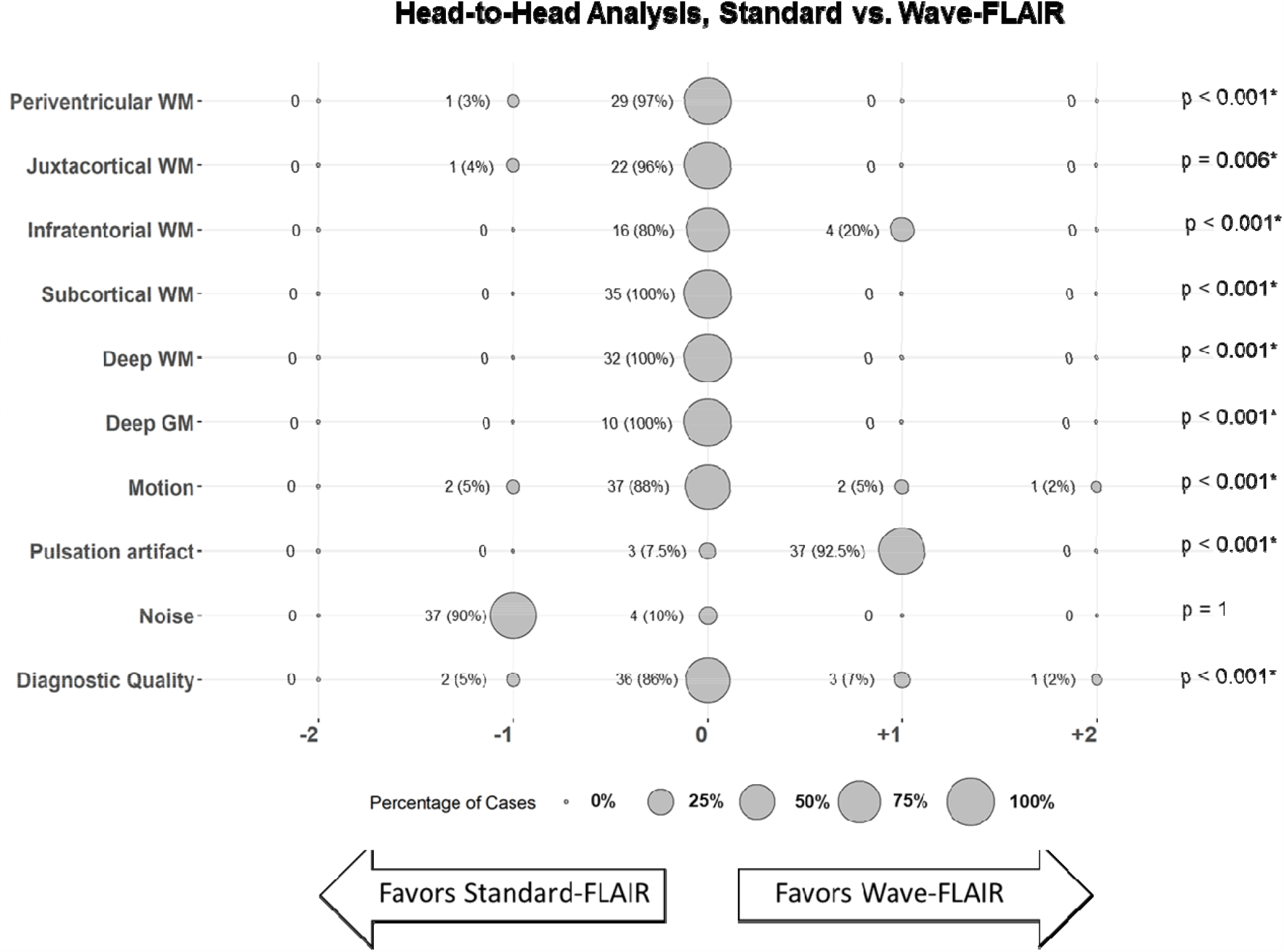
Balloon plot showing the head-to-head comparison of standard vs. Wave-FLAIR images. The size of each balloon represents the relative percentage of cases with a given score; p-values for non-inferiority testing are specified at the end of each row. The number of cases (percentage) are also noted adjacent to each balloon. ^∗^Denotes significance following correction for multiple comparisons (FDR threshold of 0.05). Raw p-values are reported in the table.

Wave-FLAIR was noninferior to standard FLAIR in terms of motion (p<0.001), with a slightly higher proportion of cases favoring Wave-FLAIR (7%) compared to standard-FLAIR (5%) (Figure 3). Wave-FLAIR demonstrated less pulsation artifact (p<0.001) in areas such as the brain stem (Figure 2e). Wave-FLAIR demonstrated more noise overall but was ultimately noninferior in overall diagnostic quality compared to standard-FLAIR (p<0.001) (Figure 3).

## Discussion

In this work, we performed a systematic quantitative evaluation of cerebral white matter lesion volumes and qualitative evaluation of lesion conspicuity, artifacts, and overall diagnostic quality of an ultrafast Wave-SPACE FLAIR sequence that was more than 2.5 times faster than the standard 3D SPACE FLAIR sequence. The results showed excellent agreement and spatial overlap between Wave and standard FLAIR white matter lesion volumes estimated by the automated segmentation tool LST. Experienced neuroradiologists rated the accelerated Wave-FLAIR images as providing equivalent visualization of lesions in the supratentorial and infratentorial white matter to the standard FLAIR images with preserved diagnostic quality. The findings support the broader application of ultrafast Wave FLAIR sequences in the evaluation of patients with white matter diseases.

White matter lesion quantification has become an increasingly important tool for characterizing the burden of disease in MS in both routine clinical evaluation as well as clinical trials.^4^ Manual white matter lesion segmentation is time-consuming and has the risk of rater bias. In addition, high image quality is needed for the best quantification. Automated lesion segmentation tools that require no or minimal training data are publicly available, including the Lesion Segmentation Toolbox (LST)^3^, LesionTOADS^21^, Salem Lesion Segmentation (SLS)^22^, and Automated Statistical Interference for Segmentation (OASIS).^23^ We chose to use LST with the lesion probability algorithm (LST-LPA)^2^ because it has high accuracy in automatically segmenting MS lesions compared to manual segmentation^7^ and requires only FLAIR images as input.^2, 7^ Here, we found comparable volumes and numbers of white matter lesions as segmented by LST on ultrafast Wave-FLAIR images compared to standard FLAIR, despite the slightly greater image noise observed in the Wave-FLAIR images. These findings were supported by the high ICC (0.99) and overall very small LVD in all brain regions (<2%) between the two sequences. The DSC were >0.9 in all supratentorial regions and were slightly lower for the infratentorial region (DSC=0.84). The lesser degree of agreement between the two sequences for infratentorial lesions likely reflects the known difficulty in detecting infratentorial lesions on FLAIR contrast images, which are overall less sensitive for posterior fossa lesions,^30, 33^ resulting in a greater difference in voxels identified as part of lesions between the two sequences. If validated in larger studies, the overall excellent agreement in lesion quantification between Wave- and standard FLAIR suggests that Wave-FLAIR could potentially replace standard FLAIR for white matter lesion quantification in clinical and research studies using imaging in MS.

In addition to volumetric measures, we also included visual evaluation of the images by multiple neuroradiologists to assess the diagnostic performance of the Wave-FLAIR sequence, which is an important part of the patient’s clinical evaluation. Wave-FLAIR provided comparable visualization of lesions in all locations to the standard sequence. The slightly greater image noise on the accelerated Wave-FLAIR images did not compromise the overall diagnostic quality. Furthermore, the Wave-FLAIR images showed reduced pulsatile flow artifact in the posterior fossa, which contributed to improved visualization of infratentorial lesions, as illustrated in Figures 2e and f. The standard FLAIR image (Figure 2e) had more pulsation artifact in the brainstem, which could lead to potential misinterpretation of a small T2 hyperintense lesion in the left lateral aspect of the pons as artifact. This lesion clearly appeared as a distinct lesion in the Wave-FLAIR image (Figure 2f) without conspicuous artifact in this area. The improved visualization of T2/FLAIR hyperintense lesions in the brainstem and cerebellum indicates that Wave-FLAIR may be useful not only in the evaluation of white matter disease burden but also in the evaluation of other conditions such as stroke and tumors.

The decreased scan time of Wave-FLAIR offers synergistic benefits for the comprehensive evaluation of white matter lesions in multiple sclerosis. Highly accelerated imaging with Wave-CAIPI has been shown to reduce motion artifact and improve the visualization of small lesions.^19^ In the current study, Wave-FLAIR was noninferior to standard-FLAIR for motion artifact. One explanation for the less pronounced improvement in motion artifact on Wave-FLAIR is that the majority of patients (85.7%) were scanned with the 20-ch coil, for which Wave-FLAIR was still 2:45 minutes in duration. We expect that motion artifact would be further reduced if more patients were scanned using the 32-ch coil (1:50 minute acquisition). The time-savings incurred by Wave-FLAIR may become more obvious when aggregated with other optimized fast 2D and 3D sequences.^12, 19, 24, 25^ For example, at our institution, we have implemented the Wave-FLAIR sequence along with optimized simultaneous multislice diffusion-weighted imaging, Wave T2 SPACE, Wave-SWI and pre- and post-contrast Wave-T1 MPRAGE sequences in the clinical multiple sclerosis brain MRI protocol, bringing the total scan time for this protocol below 20 minutes. The ability to acquire multiple 3D sequences with complementary contrasts efficiently, such as Wave-FLAIR and Wave-SWI, may encourage the greater adoption of promising imaging signs such as the central vein sign and paramagnetic rim sign^26-28^, which have greater specificity for demyelinating lesions in MS and, in the case of the paramagnetic rim sign, may have prognostic value in identifying lesions with chronic active inflammation associated with greater disability.^29^ We envision that the systematic incorporation of highly accelerated 3D Wave-CAIPI sequences into clinical MRI protocols will provide more information per unit time and enable the more comprehensive evaluation of a wide range of neurological disorders, thereby advancing clinical care and clinical research along multiple fronts. Our study had some limitations. To reduce observer bias in the qualitative evaluation, the raters were blinded to the pulse sequence, but inevitably some imaging features could help to identify the sequence and introduce observer bias. In addition, image quality also depends on the order of acquisition for each pulse sequence. In general, images acquired later in the examination would be expected to have more motion. We sought to mitigate against this bias by randomizing the acquisition order of the sequences during the study. Finally, LST underestimated total lesion numbers in patients with a high lesion load, which decreased the agreement between Wave- and standard FLAIR in certain cases, such as the assessment of confluent lesions in the periventricular area.

## Conclusion

Quantitative white matter lesion volumes and qualitative evaluation of white matter lesions imaged with an ultrafast, <3 minute Wave-FLAIR sequence showed excellent agreement with standard-FLAIR images requiring more than double the scan time in patients undergoing clinical evaluation for demyelinating disease. The findings are derived from MRI exams that were obtained as part of routine clinical work-up and/or surveillance for MS and reflect the performance of these sequences in a realistic clinical setting. The availability of ultrafast 3D sequences such as Wave-FLAIR may facilitate the more comprehensive evaluation of white matter lesions in MS and other white matter diseases.

## Data Availability

Data sharing not applicable to this article

## ABBREVIATIONS

CAIPI: Controlled Aliasing in Parallel Imaging;
SPACE: Sampling Perfection with Application optimized Contrasts by using different flip angle Evolutions;
FLAIR: Fluid Attenuated Inversion Recovery;
LST: Lesion Segmentation Tool;
MS: Multiple Sclerosis;
FSE: Fast Spin Echo;
ICC: Intra-class Correlation Coefficient;
LVD: Relative lesion Volume Difference;
DSC: Dice Similarity Coefficients;

## Acknowledgments

This work was supported by the National Institutes of Health (NIH), Grant numbers: P41-EB030006, K23-NS096056; Siemens Healthineers (research support); Massachusetts General Hospital Claflin Distinguished Scholar Award.

