## supplementary for "Evaluation of Ultrafast Wave-CAIPI 3D FLAIR in the Visualization and Volumetric Estimation of Cerebral White Matter Lesions"

**Supplementary Table 1:** Qualitive scoring criteria used for head-to-head comparison between Standard and Wave-FLAIR

| **Parameter** | **Favors image A*** | | | **Favors image B*** | |
|---|---|---|---|---|---|
| | **-1** | **-2** | **0** | **+1** | **+2** |
| **Conspicuity/visualization of lesions** | Lesions are less well visualized/conspicuous on image B and some lesions are missed | Lesions are less well visualized/conspicuous on image B but all lesions are still visualized | Equivalent | Lesions are less well visualized/conspicuous on image A but all lesions are still visualized | Lesions are less well visualized/conspicuous on image A and some lesions are missed |
| **Motion:** perceptible motion artifact when the images are optimally windowed | The B image has more motion artifacts that obscure small lesions. | The B image has more motion artifacts but it does not obscure small lesions. | Equivalent | The A image has more motion artifacts but it does not obscure small lesions. | The A image has more motion artifacts that obscures small lesions. |
| **Pulsation artifact** | Image B has more artifact and the artifact obscures underlying lesion(s). | Image B has more artifact but no lesions are obscured. | Equivalent | Image A has more artifact but no lesions are obscured. | Image A has more artifact and the artifact obscures underlying lesion(s). |
| **Noise:** perceptible noise level when the images are optimally windowed | Background noise of the B image perceptibly greater than the A image and affects the visualization of underlying structures. | Background noise of the B image perceptibly greater than the A image and does not affect the visualization of underlying structures. | Equivalent | Background noise of the A image is perceptibly greater than the B image and does not affect the visualization of underlying structures. | Background noise of the A image is perceptibly greater than the B image and affects the visualization of underlying structures. |
| **Overall diagnostic quality** | The B image has poorer image quality and the difference in quality affects the final clinical diagnosis. | The B image has poorer image quality but it does not affect the final clinical diagnosis. | Equivalent | The A image has poorer image quality but it does not affect the final clinical diagnosis. | The A image has poorer image quality and the difference in quality affects the final clinical diagnosis. |

*The Standard and Wave-FLAIR sequences were randomly positioned on either the right or left side of the screen, labeled image A and image B.

**Supplementary Table 2.** Clinical characteristics of the patients

| Characteristics | Whole cohort (n=42) | Included in Quantitative Analysis (N=36)* |
|---|---|---|
| **Female (%)** | 33 (78.6%) | 28 (77.8%) |
| **Age (year) (mean and range)** | 44.5 (23-78) | 44.8 (23-78) |
| **20-ch Coil (%)** | 36 (85.7%) | 30 (83.3%) |
| **Study indication** | | |
| **Rule out** demyelinating disease | 18 (42.9%) | 14 (38.9%) |
| Follow up of demyelinating disease | 23 (54.8%) | 21 (58.3%) |
| Other | 1 (2.4%) | 1 (2.8%) |
| **Order of the sequences** | | |
| **Standard before Wave-FLAIR (%)** | 22 (52.4%) | 17 (47.2%) |

*Six patients were excluded from the LST quantitative analysis due to absence of detectable lesions and/or severe motion artifact resulting in failure of the automated LST processing stream.

**Supplementary Table 3:** Comparison of number of lesions in brain regions between Standard and Wave-FLAIR

| | **Standard** | | **WAVE** | | **Student's t-test** |
|---|---|---|---|---|---|
| **Brain regions** | Lesions in all patients (number) | Mean (±SD) | Lesions in all patients (number) | Mean (±SD) | p-value |
| **Whole brain** | 520 | 14.4 (9.8) | 529 | 14.7 (9.7) | 0.91 |
| **Periventricular** | 223 | 6.19 (4.25) | 228 | 6.33 (4.33) | 0.89 |
| **Juxtacortical** | 79 | 2.19 (1.89) | 79 | 2.19 (1.89) | 0.99 |
| **Infra-tentorial** | 35 | 0.97 (1.4) | 35 | 0.97 (1.4) | 0.99 |
| **Deep white matter** | 99 | 2.75 (2.9) | 100 | 2.78 (2.9) | 0.97 |
| **Subcortical white matter** | 71 | 1.97 (2.47) | 72 | 2 (2.54) | 0.96 |
| **Deep gray matter** | 24 | 0.67 (1.29) | 24 | 0.67 (1.29) | 0.99 |

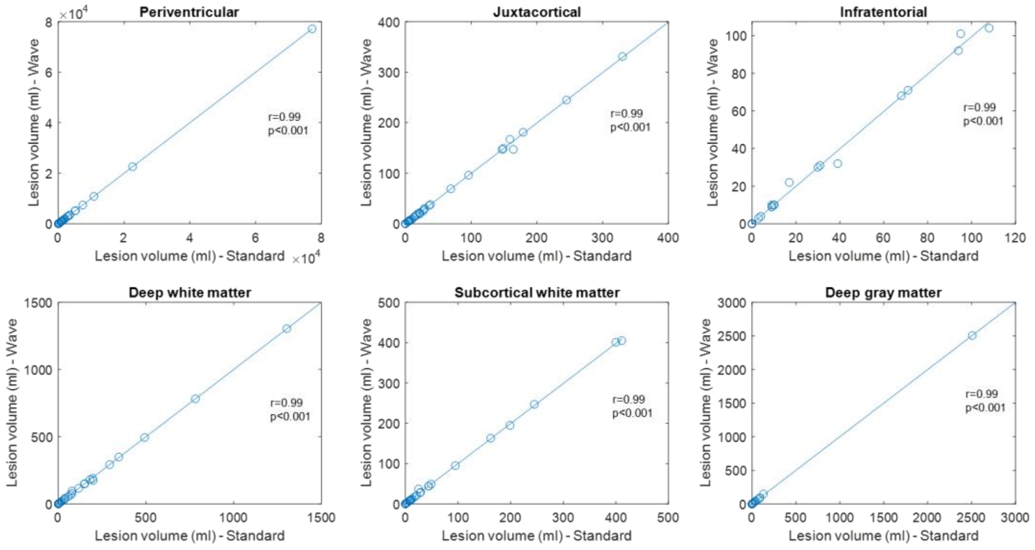


**Supplementary Figure 1.** Scatter plots of lesion volume of Standard versus Wave-FLAIR in each brain region. (r=Pearson’s correlation coefficient, p=p value)

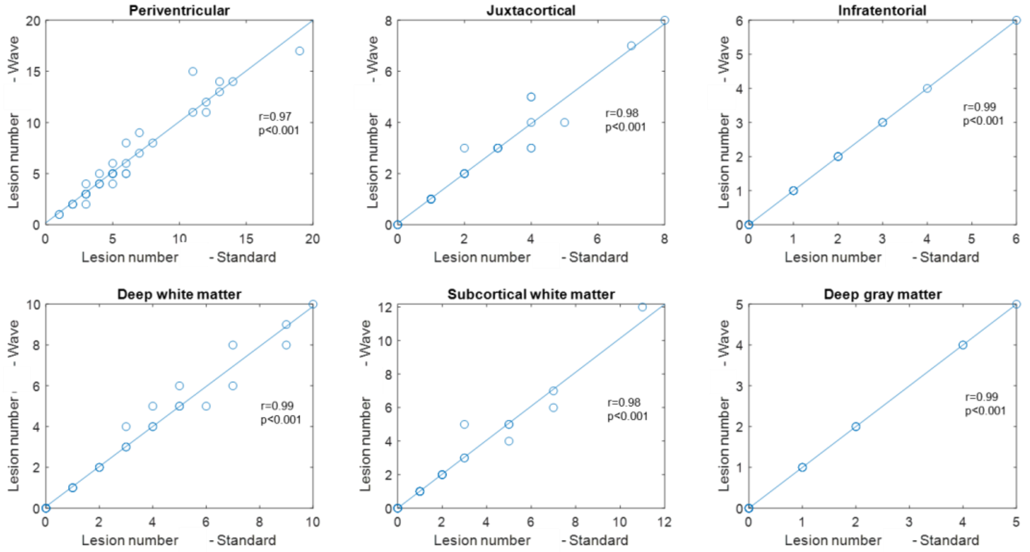


**Supplementary Figure 2.** Scatter plots of lesion number of Standard versus Wave-FLAIR in each brain region. (r=Pearson’s correlation coefficient, p=p value)
